# Cross-cancer genome-wide association study of endometrial cancer and epithelial ovarian cancer identifies genetic risk regions associated with risk of both cancers

**DOI:** 10.1101/2020.04.29.20084095

**Authors:** Dylan M. Glubb, Deborah J. Thompson, Katja K.H. Aben, Ahmad Alsulimani, Frederic Amant, Daniela Annibali, John Attia, Aurelio Barricarte, Matthias W. Beckmann, Andrew Berchuck, Marina Bermisheva, Marcus Q. Bernardini, Katharina Bischof, Line Bjorge, Clara Bodelon, Alison H. Brand, James D. Brenton, Louise Brinton, Fiona Bruinsma, Daniel D. Buchanan, Stefanie Burghaus, Ralf Butzow, Hui Cai, Michael E. Carney, Stephen J. Chanock, Chu Chen, Xiao Qing Chen, Zhihua Chen, Linda S. Cook, Julie M. Cunningham, Immaculata De Vivo, Anna deFazio, Jennifer A. Doherty, Thilo Dörk, Andreas du Bois, Alison M. Dunning, Matthias Dürst, Todd Edwards, Robert P. Edwards, Arif B. Ekici, Ailith Ewing, Peter A. Fasching, Sarah Ferguson, James M. Flanagan, Florentia Fostira, George Fountzilas, Christine M. Friedenreich, Bo Gao, Mia M. Gaudet, Jan Gawełko, Aleksandra Gentry-Maharaj, Graham G. Giles, Rosalind Glasspool, Marc T. Goodman, Jacek Gronwald, OPAL Study Group, AOCS Group, Holly R. Harris, Philipp Harter, Alexander Hein, Florian Heitz, Michelle A.T. Hildebrandt, Peter Hillemanns, Estrid Høgdall, Claus K. Høgdall, Elizabeth G. Holliday, David G. Huntsman, Tomasz Huzarski, Anna Jakubowska, Allan Jensen, Michael E. Jones, Beth Y. Karlan, Anthony Karnezis, Joseph L. Kelley, Elza Khusnutdinova, Jeffrey L. Killeen, Susanne K. Kjaer, Rüdiger Klapdor, Martin Köbel, Bozena Konopka, Irene Konstantopoulou, Reidun K. Kopperud, Madhuri Koti, Peter Kraft, Jolanta Kupryjanczyk, Diether Lambrechts, Melissa C. Larson, Loic Le Marchand, Shashikant B. Lele, Jenny Lester, Andrew J. Li, Dong Liang, Clemens Liebrich, Loren Lipworth, Jolanta Lissowska, Lingeng Lu, Karen H. Lu, Alessandra Macciotta, Amalia Mattiello, Taymaa May, Jessica McAlpine, Valerie McGuire, Iain A. McNeish, Usha Menon, Francesmary Modugno, Kirsten B. Moysich, Heli Nevanlinna, Kunle Odunsi, Håkan Olsson, Sandra Orsulic, Ana Osorio, Domenico Palli, Tjoung-Won Park-Simon, Celeste L. Pearce, Tanja Pejovic, Jennifer B. Permuth, Agnieszka Podgorska, Susan J. Ramus, Timothy R. Rebbeck, Marjorie J. Riggan, Harvey A. Risch, Joseph H. Rothstein, Ingo Runnebaum, Rodney J. Scott, Thomas A. Sellers, Janine Senz, V. Wendy Setiawan, Nadeem Siddiqui, Weiva Sieh, Beata Spiewankiewicz, Rebecca Sutphen, Anthony J. Swerdlow, Lukasz Szafron, Soo Hwang Teo, Pamela J. Thompson, Liv Cecilie Vestrheim Thomsen, Linda Titus, Alicia Tone, Rosario Tumino, Constance Turman, Adriaan Vanderstichele, Digna Velez Edwards, Ignace Vergote, Robert A. Vierkant, Zhaoming Wang, Shan Wang-Gohrke, Penelope M. Webb, Emily White, Alice S. Whittemore, Stacey J. Winham, Xifeng Wu, Anna H. Wu, Drakoulis Yannoukakos, Amanda B. Spurdle, Tracy A. O’Mara

## Abstract

Accumulating evidence suggests a relationship between endometrial cancer and epithelial ovarian cancer. For example, endometrial cancer and epithelial ovarian cancer share epidemiological risk factors and molecular features observed across histotypes are held in common (e.g. serous, endometrioid and clear cell). Independent genome-wide association studies (GWAS) for endometrial cancer and epithelial ovarian cancer have identified 16 and 27 risk regions, respectively, four of which overlap between the two cancers. Using GWAS summary statistics, we explored the shared genetic etiology between endometrial cancer and epithelial ovarian cancer. Genetic correlation analysis using LD Score regression revealed significant genetic correlation between the two cancers (*r_G_* = 0.43, P = 2.66 × 10^−5^). To identify loci associated with the risk of both cancers, we implemented a pipeline of statistical genetic analyses (i.e. inverse-variance meta-analysis, co-localization, and M-values), and performed analyses by stratified by subtype. We found seven loci associated with risk for both cancers (P_Bonferroni_ < 2.4 × 10^−9^). In addition, four novel regions at 7p22.2, 7q22.1, 9p12 and 11q13.3 were identified at a sub-genome wide threshold (P < 5 × 10^−7^). Integration with promoter-associated HiChIP chromatin loops from immortalized endometrium and epithelial ovarian cell lines, and expression quantitative trait loci (eQTL) data highlighted candidate target genes for further investigation.

## Introduction

Ovarian cancer is the eighth most commonly diagnosed cancer in women with 295,000 new cases annually^1^. Epithelial ovarian cancer accounts for ~90% of ovarian tumors and is commonly divided into five major histotypes: high-grade serous, low-grade serous, mucinous, clear cell and endometrioid^2^. Herein, “ovarian cancer” refers to epithelial types of this disease. On both histological and molecular levels, it is evident that ovarian cancer is a highly heterogeneous disease. Endometrial cancer (cancer of the uterine lining) is a comparatively understudied gynecological cancer, although it ranks fifth for cancer incidence in women globally, with 380,000 new cases diagnosed annually^1^. Endometrial cancer also has several histotypes, the most common being endometrioid (~80% of cases) but also includes serous, mucinous and clear cell.

Comparison of the epidemiology and histopathology of endometrial cancer and ovarian cancer has identified a number of similarities suggesting that shared molecular mechanisms underlie the pathology of these two diseases. Both cancers are hormone related, with epidemiological studies showing concordant direction of effect in relation to exposure to estrogen and progesterone (reviewed by Cramer ^3^). Protective factors for both types of cancer include early menopause^4,5^, late age of menarche^6,7^, longer periods of breastfeeding^8,9^, and longer use of contraceptives that include progesterone^10,11^ (i.e. factors that decrease exposure to unopposed estrogen). Although more strongly associated with endometrial cancer risk, higher body mass index (BMI) has been reported to be associated with increased risk of both cancers^12,13^.

The histotypes of endometrial cancer mirror those of ovarian cancer, albeit with varied frequencies observed across the two cancers. For example, serous histology is found in ~70% of ovarian tumors, compared with 10% of endometrial tumors, while endometrioid histology is found in ~10% of ovarian tumors and 80% of endometrial tumors. Clear cell and mucinous histologies are found in a relatively low frequency in both ovarian and endometrial tumors. Common features have been observed in similar histotypes regardless of the organ of origin. Tumors with serous histology from both the endometrium and ovary are characterized by somatic defects in the tumor suppressor gene, *TP*53^14^,^15^. Endometrioid endometrial and endometrioid ovarian tumors have both been found to contain somatic alterations in *PTEN*, *PIK3CA, ARID1A, PPP2R1A* and *CTNNB1*, although the frequencies of these mutations vary by tissue type (reviewed by McConechy, et al. ^16^). Methylation profiling has found endometrioid endometrial and endometrioid ovarian tumors cluster together^17^, and similar gene expression patterns have been observed for clear cell endometrial and clear cell ovarian tumors^18^. Further, there is increasing evidence that clear cell and endometrioid ovarian tumors arise in part from endometriosis (reviewed by King, et al. ^19^). Endometriosis is a chronic disease affecting reproductive aged women, in which endometrium grows outside of the uterus, suggesting these ovarian cancer subtypes and endometrial cancer develop from similar precursor endometrial epithelial cells.

Some, but not all germline cancer risk variants are also shared between endometrial cancer and ovarian cancer. Lynch Syndrome, characterized by germline pathogenic variants in the mismatch repair genes (i.e. *MLH1, MSH2* and *MSH6*), is associated with 40-60% and 8-15% lifetime risks of endometrial cancer and ovarian cancer, respectively^20^. Additionally, separate genome-wide association studies (GWAS) of the two cancer types have identified four genetic risk regions common to both cancers^21,22^.

Meta-analyses of GWAS datasets across etiologically-related diseases have successfully been used to increase statistical power and identify novel genetic risk regions^23,24^. Hence, in the current study, we have performed a joint meta-analysis of the largest endometrial cancer and ovarian cancer GWAS datasets to identify novel genetic loci associated with risk of both cancers, including risk variation specific to less common ovarian cancer subtypes. To identify candidate target genes at such loci, we have intersected risk variation with chromatin looping data enriched for promoter-enhancer interactions. We have also assessed associations between risk variation and gene expression to provide evidence of candidate target gene regulation and reveal further candidate genes.

## Methods

### GWAS Datasets

GWAS summary statistics were obtained from the latest meta-analyses performed by the Endometrial Cancer Association Consortium (ECAC)^21^ and the Ovarian Cancer Association Consortium (OCAC)^22^. Because of the low number of non-endometrioid endometrial cancer available in ECAC, summary statistics were provided for all endometrial cancer risk (including all endometrial cancer cases) and analyses restricted to endometrioid cases only. OCAC summary statistics were available for all ovarian cancer risk (including all ovarian cancer cases), as well analyses restricted to eight different subtypes: endometrioid histology, serous (including borderline, high- and low-grade serous cases), serous high-grade histology, serous low-grade histology, serous borderline histology, serous low-grade and borderline cases combined, clear cell histology and mucinous histology. Sample sizes for each study and subgroups analyzed are provided in **Table 1**. Details on genotyping, quality control and imputation have been previously described^21,22^. Data for approximately 10 million genetic variants (imputation quality score > 0.4 and minor allele frequency > 0.01) were available for both cancers for the present study.

**Table 1:**
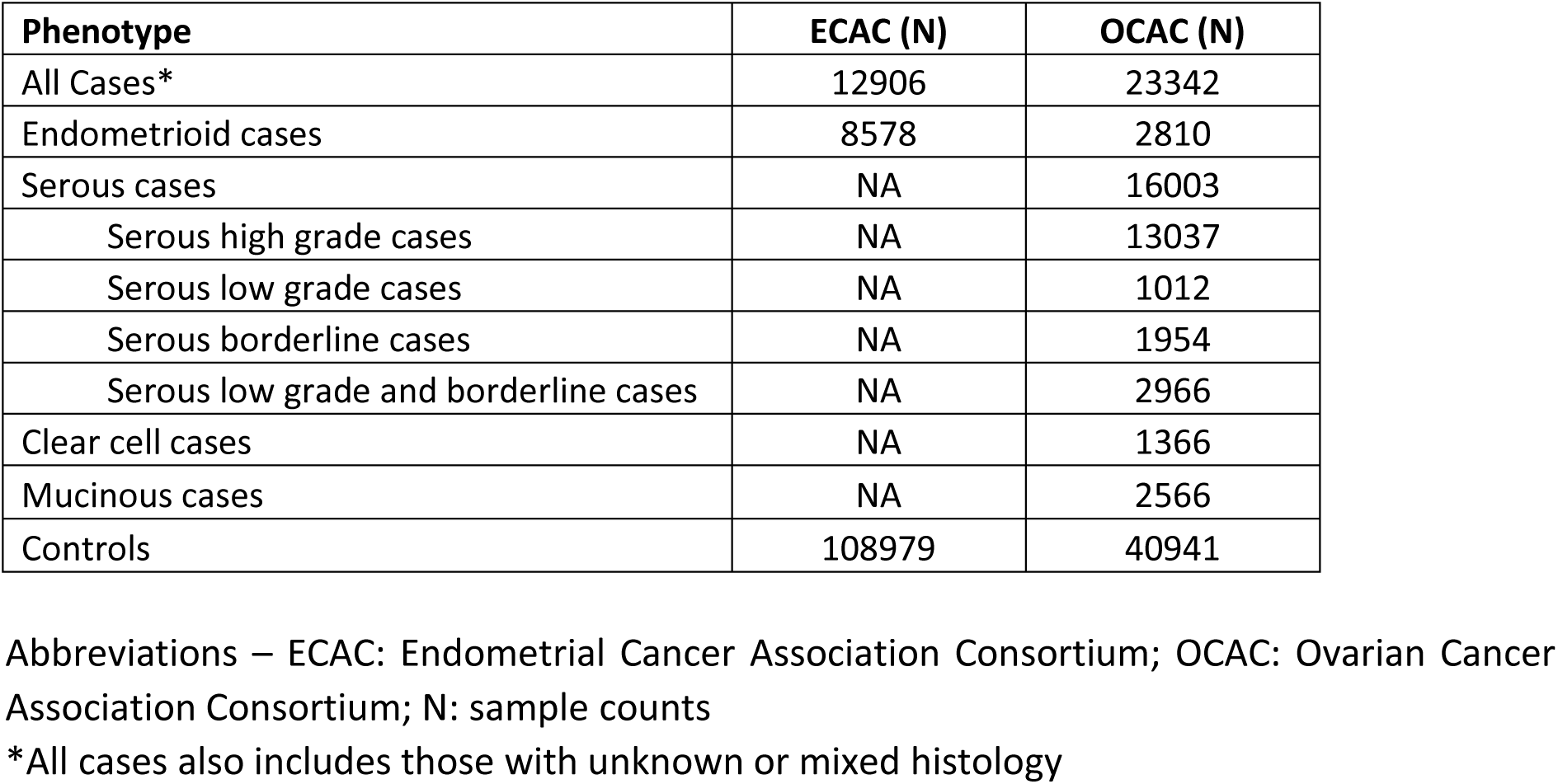
Details of samples included in the meta-analysis, by histotype

### Genetic Correlation Analyses

Genetic correlation (i.e. the estimated proportion of variance shared between two traits due to genetic factors) between endometrial cancer and ovarian cancer was assessed using linkage disequilibrium (LD) Score Regression^25^. Genetic correlation was also assessed between each of the ovarian cancer subtypes analyzed by OCAC and all endometrial cancer as well as restricted to endometrioid endometrial cancer. For this analysis, the complete set of GWAS variants were pruned to the HapMap3 variant list (~1 million variants) to provide variants with high confidence imputation scores. The major histocompatibility complex (MHC) region was removed from this analysis because of its complex LD structure.

### Cross-cancer GWAS meta-analyses

To identify joint endometrial and ovarian cancer genetic risk variants, summary statistics from ECAC and OCAC were combined by inverse-variance meta-analysis, adjusting for unknown sample overlap using MTAG^26^. Because of the significant heterogeneity in risk estimates observed for genetic variants across ovarian cancer subtypes^22^, we additionally performed meta-analysis combining results from ECAC (all endometrial cancer or endometrioid endometrial cancers) with summary statistics from each of the nine ovarian cancer subtypes analyzed by OCAC (listed in **Table 1**). To minimize false positives, following inverse-variance meta-analysis, output variants were restricted to those meeting the following criteria: (i) concordant direction of effect on risk of both cancers; (ii) no significant heterogeneity in risk estimates between the two cancers (P_het_ > 0.05); and (iii) associated with each cancer at nominal significance (P < 0.05). Counts of variants meeting these criteria are provided in **Supplementary Table S1**. M-values^27^ were generated for variants reaching with suggestive evidence of association (P < 5 x 10^−7^) using METASOFT^28^. Variants with a posterior probability for an effect in each study (M-value > 0.9) were retained for further consideration.

Loci containing variants that were statistically significant in the meta-analysis were further evaluated for co-localization by GWAS-PW^29^, using all genetic variants at the query locus. GWAS-PW estimates Bayes factors and posterior probabilities of association (PPA) for four models: (i) a locus associates with risk of endometrial cancer only; (ii) a locus associates with risk of ovarian cancer only; (iii) a locus contains a risk signal that associates with risk of both endometrial and ovarian cancers; or (iv) a locus contains two risk signals that associate independently with risk of either endometrial or ovarian cancer. Risk signals located in loci that were classified as meeting model (iii) were considered to be joint endometrial and ovarian cancer signals (PPA > 0.5).

### Cell culture

IOSE11 (immortalized ovarian surface epithelial)^30^ cells were gifted from Prof S Gayther (Cedars-Sinai Medical Center). Cells were authenticated using STR profiling and confirmed to be negative for *Mycoplasma* contamination. For routine culture, IOSE11 were grown in 1:1 MCDB105:Medium 199 with 15% FBS and antibiotics (100 IU/ml penicillin and 100 μg/ml streptomycin).

### Cell fixation

For fixation, cells (~80% confluent on 10 cm tissue culture plates) were washed with PBS and fixed at room temperature in 1% formaldehyde in PBS. After 10 min, the reaction was quenched by washing with 125 mM glycine in PBS and then adding fresh glycine-PBS. Cells were removed from the dish with a cell scraper and washed with PBS before storing cell pellets at -80°C.

### HiChIP library generation

HiChIP libraries were generated as previously^31^. Briefly, cell nuclei were extracted from fixed cell pellets and digested overnight with DpnII. After digestion, restriction fragment overhangs were filled in with biotin-dATP using the DNA polymerase I, large Klenow fragment. Proximity ligation was then performed, nuclei lysed and chromatin sheared. Sheared chromatin was incubated overnight with H3K27Ac antibody (Abcam, EP16602) to enrich for chromatin associated with promoters or enhancers. The next day Protein A beads were used to capture H3K27Ac-associated chromatin which was eluted and purified with Zymo Research concentrator columns. The DNA concentration of the purified chromatin was used to estimate the amount of TDE1 enzyme (Illumina) needed for tagmentation which was performed with biotin-labelled chromatin captured on streptavidin beads. Sequencing libraries were then generated using tagmented samples and the Nextera DNA preparation kit (Illumina). Size selection was performed using Ampure XP beads to capture 300–700 bp fragments. Two independent sequencing libraries were pooled together to provide 25 μl of library at ≥ 10 nM for Illumina HiSeq4000 (AGRF, Brisbane, QLD, Australia) paired-end sequencing with read lengths of 75 bp.

### HiChIP bioinformatics analyses

HiChIP reads (fastq files) were aligned to the human reference genome (hg19) using HiC-Pro v2.9.0^32^ and default settings were used to remove duplicate reads, assign reads to DpnII restriction fragments and filter for valid interactions as previously^31^ All valid reads from Hi-Pro were processed by the hichipper pipeline v0.7.0^33^ as previously^31^. Chromatin interactions were filtered using a minimum distance of 5 kb and a maximum of 2 Mb. The final set of chromatin loops used for further investigation were interactions that were supported by a minimum of two unique paired end tags and with a Mango^34^ q-value < 5%. Promoter-associated chromatin loops were defined as HiChIP loops with anchors within ± 3 kb of a transcription start site. Promoter-associated chromatin looping data was also available from our previous analysis of a normal immortalized endometrial cell line (E6E7hTERT)^31^.

### Credible candidate risk variants

Using 100:1 log likelihood ratios, “credible variants” (CVs) were identified at each of the joint endometrial and ovarian cancer risk regions. To identify genes that could be distally regulated by a CV, intersections of CVs with promoter-associated chromatin loops were performed using bedtools v2.28.0. Identification of genes whose expression is associated with a CV was performed by lookup of publicly available eQTL databases, including precomputed eQTL results from 336 endometrial and 318 ovarian tumors from the Cancer Genome Atlas (https://albertlab.shinyapps.io/tcga_eqtl)^35^, and from 101 non-cancerous uterus samples and 122 ovarian tissue samples from GTEx (data release v7; http://gtexportal.org)^36^. Additionally, due to the substantially increased power the sample size provided over solid tissue analyses, we accessed eQTL results from 31,684 whole blood samples (http://eqtlgen.org)^37^. Genes were considered potential targets if their expression associated with CVs that had a p-values within two orders of magnitude of the best eQTL variant in any of these eQTL datasets.

Significant genetic correlation was observed between all endometrial cancer and all ovarian cancer (*r_G_* = 0.43, P = 2.66 × 10^−5^; **Table 2**). When broken down by ovarian cancer subtype, we observed significant correlation between endometrial cancer and the following subgroups; endometrioid (*r_G_* = 0.53, P = 7.0 × 10^−3^), serous (*r_G_* = 0.42, P = 1.0 × 10^−4^) and high-grade serous ovarian cancers (*r_G_* = 0.44, P = 1.0 × 10^−4^). These correlations remained significant, although attenuated, when using endometrioid endometrial cancers only (**Table 2**).

**Table 2:** Genetic correlations between epithelial ovarian cancer subtypes and endometrial cancer (all and endometrioid) from LD score regression analysis

| Ovarian Cancer Subtype<br>(40,941 controls) | All Endometrial Cancer<br>(12,906 cases, 180,979 controls) |  | Endometrioid Endometrial Cancer<br>(8,578 cases, 46,126 controls) |  |
| --- | --- | --- | --- | --- |
| | $r_G$ (SE) | P | $r_G$ (SE) | P |
| Clear cell<br>(1,366 cases) | 0.13 (0.21) | 0.53 | 0.05 (0.23) | 0.82 |
| Endometrioid<br>(2,810 cases) | **0.53 (0.20)** | **7.00E-03** | **0.45 (0.22)** | **0.04** |
| Mucinous<br>(2,566 cases) | 0.03 (0.16) | 0.85 | -0.12 (0.18) | 0.51 |
| Serous<br>(16,003 cases) | **0.42 (0.11)** | **1.00E-04** | **0.37 (0.11)** | **9.00E-04** |
| Serous borderline<br>(1,954 cases) | 0.49 (0.56) | 0.4 | 0.68 (0.72) | 0.34 |
| Serous HG<br>(13,137 cases) | **0.44 (0.11)** | **1.00E-04** | **0.39 (0.12)** | **8.00E-04** |
| Serous LG & borderline<br>(2,966 cases) | 0.28 (0.25) | 0.25 | 0.32 (0.28) | 0.25 |
| All Ovarian<br>(23,342 cases) | **0.43 (0.10)** | **2.66E-05** | **0.36 (0.11)** | **1.40E-03** |

Abbreviations – $r_G$ : genetic correlation estimate; SE: standard error; HG: high grade. Results with a significant genetic correlation ( $P < 0.05$ ) have been bolded. The genetic heritability couldn't be estimated for one ovarian cancer subtype (serous low grade); therefore it couldn't be included in the genetic correlation analyses.

Seven genetic loci displaying evidence of a joint association with risk of both endometrial cancer (all or endometrioid histology) and ovarian cancer (all or one of the subtypes) (i.e. PPA > 0.5 for GWAS-PW model iii), passed Bonferroni-correction for multiple testing (5 × 10^−8^/17 tests = 2.9 × 10^−9^; **Table 3**). Three of these loci belong to regions that have previously been reported as being associated with risk of both cancers (8q24, 17q12 and 17q21.32), although the 17q21.32 region had not been reported to be associated with the specific subtypes of ovarian cancer found in this meta-analysis (**Table 3**). One of the seven loci (2p16.1) has been previously reported as being associated with risk of endometrial cancer, but not with ovarian cancer risk. The three remaining loci (5p15.33, 9q34.2 and 10p12.31) have been previously reported as associated with risk of all ovarian cancer and serous ovarian cancer but not with endometrial cancer risk below GWAS significance levels; however, associations between endometrial cancer and variants in the 5p15.33 (*TERT*) region have been reported in a candidate-region study^38^. Additionally, we identified four novel loci with sub-GWAS significance levels (P < 5 × 10^−7^) that had not been previously reported as being associated with risk of either cancer at genome-wide levels of significance (7p22.2, 7q22.1, 9p12 and 11q13.3, **Figure 1**).

**Table 3:** Results from GWAS meta-analysis of endometrial cancer and epithelial ovarian cancer

|  |  |  |  |  |  |  |  | Endometrial Cancer |  |  | Ovarian Cancer |  |  | Meta-analysis |  |  |
| --- | --- | --- | --- | --- | --- | --- | --- | --- | --- | --- | --- | --- | --- | --- | --- | --- |
| Region | ECAC Phenotype | OCAC Phenotype | Lead Variant | Chr:Pos (hg19) | EA/OA | Freq EA (ECAC/OCAC) | OncoArray INFO Score (ECAC/OCAC) | OR (95% CI) | P-value | M-value | OR (95% CI) | P-value | M-value | OR (95% CI) | P-value | Model 3 PPA |
| **<u>Known endometrial and ovarian cancer risk regions</u>** |  |  |  |  |  |  |  |  |  |  |  |  |  |  |  |  |
| 8q24.21 | endometrioid | all | rs10103314 | 8:129560744 | C/A | 0.13/0.13 | 1.00/0.99 | 0.86 (0.82-0.91) | 9.05E-08 | 1.00 | 0.85 (0.82-0.88) | 4.91E-16 | 1.00 | 0.85 (0.82-0.88) | 1.49E-20 | 0.90 |
| 17q12 | all | clear cell | rs11263763 | 17:36103565 | A/G | 0.55/0.52 | 1.00/1.00 | 1.15 (1.12-1.19) | 4.01E-20 | 1.00 | 1.25 (1.15-1.35) | 3.46E-08 | 1.00 | 1.16 (1.13-1.2) | 2.46E-24 | 1.00 |
| 17q12 | endometrioid | clear cell | rs11263763 | 17:36103565 | A/G | 0.55/0.52 | 1.00/1.00 | 1.15 (1.11-1.19) | 1.23E-14 | 1.00 | 1.25 (1.15-1.35) | 3.46E-08 | 1.00 | 1.17 (1.13-1.21) | 2.20E-19 | 1.00 |
| 17q21.32 | all | clear cell | rs882380 | 17:46294236 | A/C | 0.61/0.60 | 0.99/0.97 | 1.10 (1.06-1.13) | 4.66E-09 | 1.00 | 1.09 (1.00-1.18) | 0.04 | 0.94 | 1.10 (1.06-1.13) | 1.91E-09 | 0.85 |
| 17q21.32 | endometrioid | clear cell | rs882380 | 17:46294236 | A/C | 0.61/0.60 | 0.99/0.97 | 1.11 (1.07-1.15) | 1.25E-08 | 1.00 | 1.09 (1.00-1.18) | 0.04 | 0.94 | 1.11 (1.07-1.15) | 4.67E-09 | 0.90 |
| 17q21.32 | all | endometrioid | rs882380 | 17:46294236 | A/C | 0.61/0.60 | 0.99/0.97 | 1.10 (1.06-1.13) | 4.66E-09 | 1.00 | 1.09 (1.03-1.15) | 3.44E-03 | 0.99 | 1.09 (1.06-1.13) | 2.90E-10 | 0.91 |
| 17q21.32 | endometrioid | endometrioid | rs882380 | 17:46294236 | A/C | 0.61/0.60 | 0.99/0.97 | 1.11 (1.07-1.15) | 1.25E-08 | 1.00 | 1.09 (1.03-1.15) | 3.44E-03 | 0.99 | 1.11 (1.07-1.14) | 6.91E-10 | 1.00 |
| 17q21.32 | all | serous borderline | rs12950225 | 17:46145200 | G/A | 0.58/0.57 | 1.00/1.00 | 1.08 (1.05-1.12) | 1.98E-07 | 1.00 | 1.10 (1.03-1.18) | 5.64E-03 | 0.99 | 1.09 (1.06-1.12) | 1.26E-08 | 1.00 |
| 17q21.32 | endometrioid | serous borderline | rs882380 | 17:46294236 | A/C | 0.61/0.60 | 0.99/0.97 | 1.11 (1.07-1.15) | 1.25E-08 | 1.00 | 1.15 (1.07-1.23) | 9.56E-05 | 1.00 | 1.12 (1.08-1.16) | 2.88E-11 | 0.99 |
| 17q21.32 | all | serous LG & borderline | rs882380 | 17:46294236 | A/C | 0.61/0.60 | 0.99/0.97 | 1.10 (1.06-1.13) | 4.66E-09 | 1.00 | 1.14 (1.08-1.21) | 6.73E-06 | 1.00 | 1.11 (1.08-1.14) | 3.10E-12 | 0.98 |
| 17q21.32 | all | serous LG | rs882380 | 17:46294236 | A/C | 0.61/0.60 | 0.99/0.97 | 1.10 (1.06-1.13) | 4.66E-09 | 1.00 | 1.12 (1.02-1.23) | 0.02 | 0.96 | 1.10 (1.07-1.13) | 1.84E-09 | 0.99 |
| **<u>Known endometrial cancer risk regions</u>** |  |  |  |  |  |  |  |  |  |  |  |  |  |  |  |  |
| 2p16.1 | all | clear cell | rs148261157 | 2:60897579 | A/G | 0.04/0.04 | 0.89/0.87 | 1.26 (1.16-1.36) | 3.39E-08 | 1.00 | 1.37 (1.11-1.69) | 3.18E-03 | 0.99 | 1.27 (1.18-2.78) | 1.85E-09 | 0.96 |
| 2p16.1 | endometrioid | clear cell | rs7579014 | 2:60707894 | A/G | 0.64/0.63 | 0.99/0.98 | 1.10 (1.06-1.14) | 6.16E-07 | 1.00 | 1.13 (1.04-1.22) | 4.24E-03 | 0.99 | 1.10 (1.07-1.57) | 2.92E-08 | 0.71 |
| **<u>Known ovarian cancer risk regions</u>** |  |  |  |  |  |  |  |  |  |  |  |  |  |  |  |  |
| 5p15.33 | all | all | rs7725218 | 5:1282414 | A/G | 0.36/0.35 | 0.97/0.94 | 1.07 (1.04-1.11) | 1.12E-05 | 1.00 | 1.10 (1.07-1.13) | 1.76E-11 | 1.00 | 1.09 (1.07-1.11) | 2.71E-14 | 1.00 |
| 5p15.33 | endometrioid | all | rs7726159 | 5:1282319 | A/C | 0.34/0.34 | 0.98/0.94 | 1.08 (1.04-1.12) | 7.90E-05 | 1.00 | 1.10 (1.07-1.13) | 1.04E-11 | 1.00 | 1.09 (1.07-1.12) | 5.23E-14 | 1.00 |
| 5p15.33 | all | serous | rs6897196 | 5:1280938 | G/A | 0.40/0.39 | 1.00/0.98 | 1.07 (1.03-1.10) | 6.46E-05 | 0.99 | 1.11 (1.08-1.15) | 2.07E-11 | 1.00 | 1.09 (1.07-1.12) | 2.21E-13 | 1.00 |
| Region | ECAC Phenotype | OCAC Phenotype | Lead Variant | Chr:Pos (hg19) | EA/OA | Freq EA (ECAC/OCAC) | OncoArray INFO Score (ECAC/OCAC) | OR (95% CI) | P-value | M-value | OR (95% CI) | P-value | M-value | OR (95% CI) | P-value | Model 3 PPA |
| 5p15.33 | endometrioid | serous | rs7725218 | 5:1282414 | A/G | 0.36/0.35 | 0.97/0.94 | 1.08<br>(1.04-1.12) | 6.12E-05 | 0.99 | 1.13<br>(1.09-1.16) | 1.5E-13 | 1.00 | 1.11<br>(1.08-1.14) | 1.40E-15 | 1.00 |
| 5p15.33 | all | serous HG | rs7725218 | 5:1282414 | A/G | 0.36/0.35 | 0.97/0.94 | 1.07<br>(1.04-1.11) | 1.12E-05 | 1.00 | 1.12<br>(1.09-1.16) | 4.4E-12 | 1.00 | 1.10<br>(1.07-1.12) | 1.07E-14 | 1.00 |
| 5p15.33 | endometrioid | serous HG | rs7725218 | 5:1282414 | A/G | 0.36/0.35 | 0.97/0.94 | 1.08<br>(1.04-1.12) | 6.12E-05 | 0.99 | 1.12<br>(1.09-1.16) | 4.4E-12 | 1.00 | 1.10<br>(1.08-1.13) | 2.34E-14 | 1.00 |
| 5p15.33 | all | serous LG & borderline | rs2853672 | 5:1292983 | A/C | 0.48/0.49 | 1.00/1.00 | 0.94<br>(0.91-0.96) | 1.30E-05 | 1.00 | 0.88<br>(0.83-0.93) | 5.72E-06 | 1.00 | 0.92<br>(0.90-0.95) | 1.03E-08 | 1.00 |
| 5p15.33 | endometrioid | serous LG & borderline | rs2853672 | 5:1292983 | A/C | 0.48/0.49 | 1.00/1.00 | 0.93<br>(0.89-0.96) | 2.27E-05 | 1.00 | 0.88<br>(0.83-0.93) | 5.72E-06 | 1.00 | 0.91<br>(0.88-0.94) | 7.20E-09 | 1.00 |
| 9q34.2 | all | all | rs635634 | 9:136155000 | T/C | 0.20/0.20 | 1.00/1.00 | 1.06<br>(1.02-1.10) | 1.48E-03 | 0.99 | 1.10<br>(1.07-1.14) | 3.08E-09 | 1.00 | 1.09<br>(1.06-1.11) | 3.46E-10 | 0.91 |
| 9q34.2 | all | serous | rs687289 | 9:136137106 | A/G | 0.35/0.34 | 1.00/1.00 | 1.07<br>(1.03-1.10) | 6.39E-05 | 1.00 | 1.10<br>(1.06-1.13) | 1.35E-08 | 1.00 | 1.08<br>(1.06-1.11) | 4.15E-11 | 0.80 |
| 10p12.31 | all | all | rs564819152 | 10:21820650 | G/A | 0.32/0.32 | 1.00/0.97 | 1.05<br>(1.02-1.08) | 2.55E-03 | 0.97 | 1.09<br>(1.06-1.12) | 2.52E-10 | 1.00 | 1.08<br>(1.05-1.10) | 8.73E-11 | 0.99 |
| 10p12.31 | all | serous | rs7090708 | 10:21929179 | G/A | 0.33/0.33 | 1.00/0.99 | 1.05<br>(1.02-1.08) | 2.6E-03 | 0.96 | 1.09<br>(1.06-1.13) | 1.92E-08 | 1.00 | 1.07<br>(1.05-1.10) | 3.62E-09 | 0.99 |
| 10p12.31 | all | serous HG | rs7090708 | 10:21929179 | G/A | 0.33/0.33 | 1.00/0.99 | 1.05<br>(1.02-1.08) | 2.6E-03 | 0.92 | 1.10<br>(1.07-1.14) | 5.02E-08 | 1.00 | 1.07<br>(1.05-1.10) | 7.63E-09 | 0.99 |
| **Novel regions** |  |  |  |  |  |  |  |  |  |  |  |  |  |  |  |  |
| 7p22.2 | all | all | rs13221982 | 7:3865621 | C/T | 0.06/0.06 | 0.98/0.98 | 1.13<br>(1.06-1.21) | 1.32E-04 | 1.00 | 1.12<br>(1.06-1.18) | 6.85E-05 | 1.00 | 1.13<br>(1.08-1.18) | 1.57E-07 | 0.90 |
| 9p12 | endometrioid | serous LG & borderline | rs2475339 | 9:10262484 | T/C | 0.83/0.83 | 0.99/0.99 | 0.89<br>(0.85-0.93) | 8.64E-07 | 1.00 | 0.90<br>(0.84-0.97) | 4.50E-03 | 0.99 | 0.89<br>(0.86-0.93) | 4.36E-08 | 0.94 |
| 7q22.1 | all | serous borderline | rs139380031 | 7:98911827 | A/C | 0.03/0.03 | 0.97/0.95 | 0.77<br>(0.70-0.85) | 5.98E-07 | 1.00 | 0.77<br>(0.61-0.97) | 0.03 | 0.95 | 0.77<br>(0.70-0.85) | 1.28E-07 | 0.57 |
| 11q13.3 | endometrioid | all | rs7118966 | 11:69019272 | C/T | 0.24/0.25 | 1.00/1.00 | 0.93<br>(0.89-0.97) | 4.30E-04 | 0.99 | 0.94<br>(0.91-0.97) | 1.96E-05 | 1.00 | 0.93<br>(0.91-0.96) | 1.25E-07 | 0.82 |

Abbreviations – EA: Effect Allele; OA: Other Allele; EAF: Effect Allele Frequency; OR: Odds Ratio; CI: Confidence Interval; PPA: Posterior Probability of Association; HG: High grade; LG: Low grade Italicized results meet suggestive association ( $P < 5 \times 10^{-7}$ )

**Figure 1.**
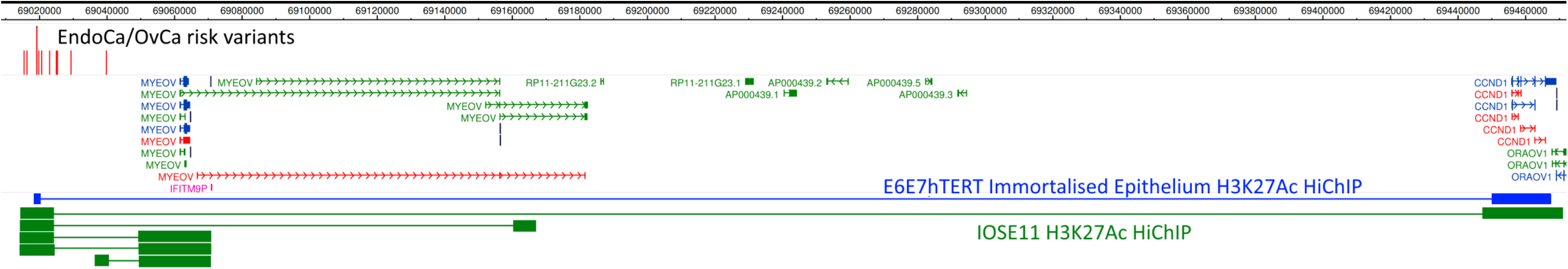
Promoter-associated chromatin looping by HiChIP identifies candidate target genes at the 11q13.3 locus. Promoter-associated loops were intersected with joint endometrial and ovarian cancer risk CVs (colored red), revealing chromatin loops that interact with the promoter of *CCDN1* in both an immortalized endometrium epithelial cell line (E6E7hTERT, colored blue) and an immortalized ovarian surface epithelial cell line (IOSE11, colored green).

We identified a total of 22 candidate target genes at the 11 identified joint endometrial and ovarian cancer risk loci using a number of approaches (**Table 4, Supplementary Table S2**). Log likelihood ratios identified a median of 20 CVs per locus (range 1–73, **Supplementary Table S3**). Using H3K27Ac-associated chromatin looping data from normal immortalized ovarian surface epithelial cells and the same data previously generated from a normal immortalized endometrium cell line^31^, we intersected CVs coincident with putative enhancers (marked by H3K27Ac) belonging to promoter-associated loops. We found looping between such enhancers and the promoters of 14 genes (at five of the 11 loci) to be common to both immortalized endometrium and ovarian surface epithelial cell lines (e.g. **Figure 1**). These included genes which encode proteins involved in relevant processes such as steroid hormone metabolism (*CYP3A43*), estrogen response (*GPER*) and oncogenesis (*MYC, CCDN1*). Four of the 14 candidate target genes identified by chromatin looping also had CV located in the promoter, indicating potential to regulate expression (**Table 4**). An additional five genes were identified as candidate targets with CVs located in the corresponding promoters (**Table 4**). Interrogation of five relevant public eQTL databases revealed CVs to be associated with the expression of four genes (*ABO, BCL11A, HOXB2* and *SNX11*), highlighting them as candidate targets. One of these, *SNX11*, had also been identified through the chromatin looping analyses and a CV was located in its promoter. Notably, we observed that increased expression of *ABO* associated with risk allele of CVs at the 9q34.2 locus in all five eQTL datasets: blood, non-cancerous uterine and ovarian tissues, and endometrial and ovarian tumors.

**Table 4:**
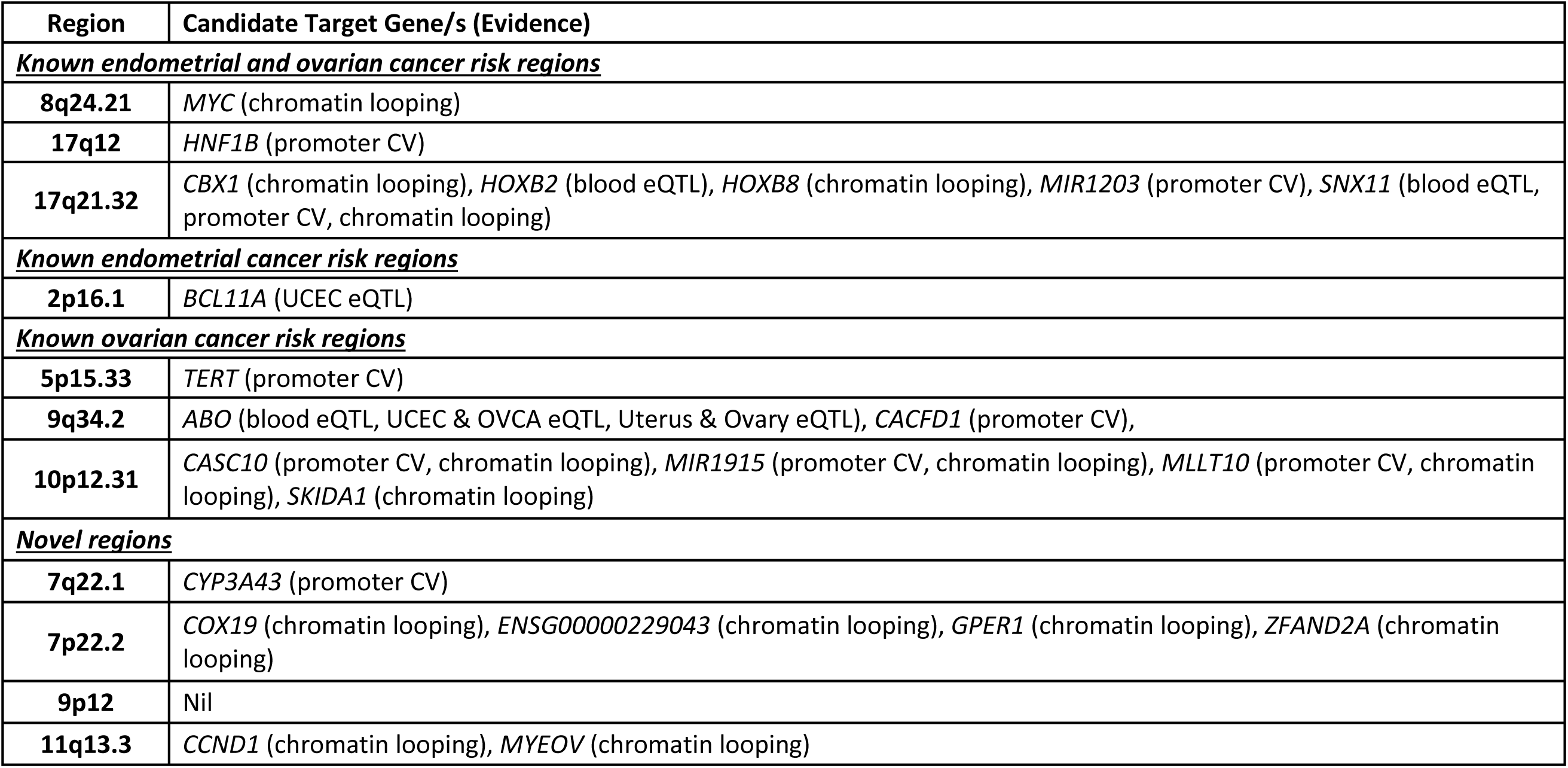
Candidate target genes at joint endometrial cancer and epithelial ovarian cancer risk loci.

| Region | Candidate Target Gene/s (Evidence) |
| --- | --- |
| **<u>Known endometrial and ovarian cancer risk regions</u>** |  |
| **8q24.21** | *MYC* (chromatin looping) |
| **17q12** | *HNF1B* (promoter CV) |
| **17q21.32** | *CBX1* (chromatin looping), *HOXB2* (blood eQTL), *HOXB8* (chromatin looping), *MIR1203* (promoter CV), *SNX11* (blood eQTL, promoter CV, chromatin looping) |
| **<u>Known endometrial cancer risk regions</u>** |  |
| **2p16.1** | *BCL11A* (UCEC eQTL) |
| **<u>Known ovarian cancer risk regions</u>** |  |
| **5p15.33** | *TERT* (promoter CV) |
| **9q34.2** | *ABO* (blood eQTL, UCEC & OVCA eQTL, Uterus & Ovary eQTL), *CACFD1* (promoter CV), |
| **10p12.31** | *CASC10* (promoter CV, chromatin looping), *MIR1915* (promoter CV, chromatin looping), *MLLT10* (promoter CV, chromatin looping), *SKIDA1* (chromatin looping) |
| **<u>Novel regions</u>** |  |
| **7q22.1** | *CYP3A43* (promoter CV) |
| **7p22.2** | *COX19* (chromatin looping), *ENSG00000229043* (chromatin looping), *GPER1* (chromatin looping), *ZFAND2A* (chromatin looping) |
| **9p12** | Nil |
| **11q13.3** | *CCND1* (chromatin looping), *MYEOV* (chromatin looping) |

## Discussion

In this study, we have performed the first cross-cancer GWAS analysis of endometrial cancer and ovarian cancer. Genetic correlation analyses found significant correlation between the two cancers, particularly between all endometrial cancer (and its endometrioid subtype) and the serous (high- and low-grade combined) or endometrioid ovarian cancer subtypes. Our pipeline of genetic analyses, stratifying by subtype, allowed us to identify seven joint endometrial cancer and ovarian cancer genetic risk loci. Three of these loci were located in regions that had been previously associated with both cancers, one was located in a known endometrial cancer risk region and the remaining three were located in known ovarian cancer risk regions. Four novel genetic risk loci for these two cancers did not reach the statistical threshold for significance but were highlighted as of potential interest, requiring further study to confirm their status.

Joint endometrial and ovarian cancer risk loci are located in the 8q24.21 and 5p15.33 regions, previously described as “cancer GWAS nexus regions”^39^ since genetic variation at these regions has been associated with many different types of cancer. 8q24.21 has been previously identified as a genetic risk region for both endometrial cancer and ovarian cancer^21,22^. CVs in a putative enhancer at the 8q24.21 joint endometrial and ovarian cancer risk locus showed evidence of chromatin looping to the promoter of the pan-cancer *MYC* oncogene in immortalized endometrial epithelial and ovarian surface epithelial cell lines. A previous study of the 5p15.33 multi-cancer risk region, containing the *TERT* gene, identified two independent signals for ovarian cancer risk: one (lead variant rs7705526) associated with serous borderline ovarian cancer risk and the other (lead variant rs10069690) associated with serous invasive ovarian cancer risk^40^. Although not previously associated with risk of endometrial cancer at genome-wide significance, a candidate fine-mapping study of 5p15.33 did highlight three independent endometrial cancer risk signals at this locus at study-wide significance^38^, one of which was shared with the serous borderline ovarian cancer risk signal. The present analysis identified this signal as a joint endometrial and ovarian cancer risk signal, with CVs in the *TERT* promoter highlighting this gene as a likely target. Moreover, the TERT protein has been heavily implicated in cancer development (reviewed in Yuan, et al. ^41^) and has oncogenic interactions with MYC (reviewed in Pestana, et al. ^42^).

Our results suggest, at a sub-genome wide significance level, a potential joint endometrial and ovarian cancer risk signal at another cancer GWAS nexus region, 11q13.3. Originally identified as a prostate cancer risk locus, 11q13.3 also contains risk signals for melanoma, breast cancer and renal cancer (https://www.ebi.ac.uk/gwas/). Although the results from the present study require validation, the identification of a shared endometrial and ovarian cancer risk signal at 11q13.3 would provide further evidence that this region is important for cancer development. At this locus, chromatin looping data showed that CVs in a putative enhancer looped to the promoters of *MYEOV* and *CCND1*, in immortalized endometrial epithelial and ovarian surface epithelial cell lines. *CCND1* (encoding cyclin D1) is of particular interest as it is frequently amplified in human cancers and has been identified as a pan-cancer driver gene^43^. Cyclin D1 is considered an oncogene due to its central role in cell cycle regulation, and ability to promote cell proliferation^44^. *CCND1* has been found to be significantly mutated in gynecological (endometrial, ovarian and cervical cancer, and uterine carcinosarcoma) and breast cancers^45^. The results of our genetic association analyses and integrative analyses of chromatin interactions results provide additional support that *CCND1* is important in the development of endometrial cancer and ovarian cancer.

Our analysis identified the 17q12 region as a joint endometrial and ovarian cancer risk region, associating with clear cell ovarian cancer. The 17q12 region, containing *HNF1B*, has been previously associated with risk of endometrial cancer and ovarian cancer^46–49^. Significant heterogeneity in risk estimates has been observed across ovarian cancer histotypes at this locus. The minor allele of the lead ovarian cancer risk variant previously identified at this region associated with increased serous (high- and low-grade combined) ovarian cancer risk but decreased clear cell ovarian cancer risk^48,49^. Further genotyping had resolved this region into two risk signals for ovarian cancer risk: one in intron 1 of *HNF1B* for clear cell ovarian cancer risk (rs11651775; the same signal for endometrial cancer risk) and another in intron 3 for serous ovarian cancer risk (rs7405776)^49^. Our results confirm that joint endometrial and ovarian cancer risk variants at 17q12 map to the same signal as that for that previously reported for endometrial cancer and the clear cell ovarian subtype. *HNF1B* is a likely target of endometrial and ovarian cancer risk variation, with CVs located in the promoter region of this gene. We have previously demonstrated that these variants affect activity of the *HNF1B* promoter^46^, which may lead to increased secretion of insulin, a risk factor for endometrial cancer^50^.

The 17q21.32 region is a known shared endometrial^21^ and ovarian cancer^22^ risk region. The joint endometrial and ovarian cancer signal found in the present study (lead SNP rs882380) is the same as that previously identified for endometrial cancer, but is independent of the signal previously found for all invasive and high-grade serous ovarian cancer risk (lead SNP rs7207826, r^2^ = 0.06 with rs882380). The joint endometrial and ovarian cancer signal associates specifically with risk of clear cell, endometrioid, serous low-grade, serous low-grade and borderline combined, and serous borderline ovarian cancer subtypes. Clear cell, endometrioid and serous low-grade ovarian cancers are often referred to as endometriosis-associated ovarian cancers due to the increased risk of these ovarian cancer subtypes with endometriosis^51^. Epidemiological and molecular data provide strong evidence that clear cell and endometrioid ovarian cancer arise in part from endometriosis (reviewed by King, et al. ^19^). The joint endometrial and ovarian cancer signal identified in the present study at 17q21.32 was also found in a joint GWAS analysis of endometrial cancer and endometriosis^52^, and subsequently found to be associated with endometriosis risk independently^53^. Five candidate target genes were identified at this locus, all of which we had previously found to be candidate targets of the original endometrial cancer signal through chromatin looping studies of endometrial cancer cell lines^31^.

Another potential joint endometrial and ovarian cancer signal, 9p12, associated with risk of serous low-grade ovarian cancer, has also been previously identified as a joint endometrial cancer and endometriosis risk locus^52^. These findings at 17q21.32 and 9p12, add to the body of evidence for the relationship between endometriosis and specific ovarian cancer subtypes^19,51^, and provide further support for shared genetic etiology between endometriosis and endometrial cancer^52^. CVs at the 9p12 joint risk locus were located intronic to *PTPRD*, but no candidate target genes were identified through chromatin looping or eQTL analyses. PTPRD protein is involved in the STAT3 pathway which has been implicated as a potential target for both endometrial cancer^54^ and ovarian cancer^55^.

The 2p16.1 region is a known endometrial cancer risk locus and was found to associate with the risk of clear cell ovarian cancer only. Interestingly, we previously found evidence that this locus may have a stronger association with risk of non-endometrioid endometrial cancer, with the strongest effect observed for clear cell endometrial cancer subtype (128 cases and 26,638 controls; rs148261157 OR 2.36; 95% CI 1.07 - 5.19)^21^. *BCL11A* was identified as a candidate target gene through eQTL analysis of endometrial tumors. We had previously found that *BCL11A* was a candidate target gene at the endometrial cancer risk locus through chromatin looping studies in endometrial cancer cells^31^. The eQTL finding suggested that reduced expression of *BCL11A* may increase endometrial/clear cell ovarian cancer risk. Indeed, some studies have shown that *BCL11A* acts as a proto-oncogene^56,57^; however, others suggest that overexpression of *BCL11A* results in anti-cancer effects^58^. Notably, *BCL11A* has been found to be mutated in clear cell ovarian cancer^59,60^, providing further evidence that the expression of *BCL11A* explains, at least in part, the mechanism underlying the risk association with both endometrial cancer and clear cell ovarian cancer.

The 9q34.2 region is a known ovarian cancer risk locus that is highly pleiotropic, having been previously associated with gastric and pancreatic cancers, in addition to a wide range of traits including blood cell counts, the tumor marker CEA (carcinoembryonic antigen), circulating cholesterol, bone mineral density and levels of proteins related to angiogenesis (e.g. VEGFR-2 and angiopoietin)(https://www.ebi.ac.uk/gwas/). eQTL data from normal, tumor endometrial and ovarian tissue, as well as blood, provide evidence that *ABO* is a regulatory target of CVs at this locus. *ABO* encodes an enzyme with glycosyltransferase activity and determines human ABO blood group antigens. It is not immediately apparent how *ABO* may mediate cancer risk but its encoded glycosyltransferase can affect cell recognition and adhesion, and activation of T and natural killer cells (reviewed by Arend ^61^).

The 10p12.31 region is another known ovarian cancer risk locus that is also pleiotropic, having been previously associated with breast cancer as well as with traits related to obesity such as BMI, body fat percentage and physical activity (https://www.ebi.ac.uk/gwas/). *MLLT10* was identified as a candidate target gene at this locus, through chromatin looping analysis and localization of a CV to its promoter, and has been found to be a partner gene for chromosomal rearrangements that result in leukaemia^62^. Another biologically relevant candidate target gene at this locus is *MIR1915* whose expression is upregulated by p53 in response to DNA damage, subsequently leading to increased apoptosis^63^.

Two of the sub-genome wide significant endometrial/ovarian cancer risk regions (7q22.1 and 7p22.2) may relate to circulating hormone levels or regulation. At 7q22.1, GWAS have previously revealed associations with androgen and progesterone levels^64^. The sole candidate target gene at this locus, *CYP3A43*, encodes a cytochrome P450 enzyme that may be involved in androgen metabolism^65^ and is upregulated in ovarian tumors^66^. At 7p22.2, the candidate target gene *GPER1*, identified through chromatin looping, encodes an estrogen receptor that induces endometrial and ovarian cancer cell proliferation in response to estrogen (reviewed in Prossnitz and Barton ^67^). Further, it appears that androgen can also bind to GPER1 to stimulate cancer cell growth^68^.

Despite these findings, the present study does have some limitations. The low numbers of non-endometrioid endometrial cancers meant we could not explore the relationship of these endometrioid histotypes and their relationship with ovarian cancer. Another limitation was the use of cell lines to model chromatin looping that occurs in endometrial and ovarian tissue, with chromatin looping potentially impacted by the immortalization and 2D-culturing processes of cell lines, or mutations gained through routine passaging of cells. Only one endometrial and one ovarian cell line were used to identify chromatin looping events. These experiments should be repeated in additional endometrial and ovarian cell lines, representing tumor subtypes. One of the four regions previously identified to be associated with both cancers, located at 1p34, was not identified in the present analysis. This locus was originally found in a combined analysis of the OCAC with a cohort of *BRCA1/2* carriers with ovarian cancer, i.e. the Consortium of Investigators of Modifiers of BRCA1 and BRCA2 (CIMBA)^69^. The CIMBA study was not included in the present study, perhaps explaining why it was not identified in our analysis as a joint endometrial and ovarian cancer locus. Future analysis of this region, in the context of *BRCA1/2* carrier status will be required to explore how this region affects endometrial cancer and ovarian cancer risk.

In summary, using endometrial and ovarian cancer GWAS summary statistics we have been able to identify seven joint risk loci for these cancers, with an additional four novel potential risk regions at a sub-GWAS significance level. Further studies are required to validate these findings in larger sample sets. Notably, we also found significant genetic correlation between the two cancers, supported by the observed epidemiological and histopathological similarities. These findings support the need for larger GWAS of endometrial and ovarian cancer, in particular focusing on their minor subtypes to further explore shared genetic etiology. Integration of CVs with chromatin looping and eQTL data has identified several plausible candidate target genes, including those at potentially novel risk loci. Although the role of these genes in endometrial and ovarian cancer development should be explored in future studies, the current findings provide insights into the shared biology of endometrial and ovarian cancer.

## Data Availability

OCAC and ECAC summary statistics are available from the GWAS catalog. The E6E7hTERT HiChIP data are available from GEO and IOSE11 HiChIP data will be made available upon publication.

https://www.ebi.ac.uk/gwas/

## Acknowledgements

We thank ECAC and OCAC for provision of summary statistics to perform this study. We thank Siddhartha Kar for his helpful discussions and advice for designing the genetic analysis approaches. Full acknowledgements and funding for ECAC and OCAC can be found in the Supplementary Note.

## Notes

**Financial Support:** TAO’M is supported by a National Health and Medical Research Council (NHMRC) Early Career Fellowship (APP1111246) and Investigator Fellowship (APP1173170), ABS is supported by an NHMRC Senior Research Fellowship (APP1061779) and Investigator Fellowship (APP1177524). This work was supported by a Cancer Australia PdCCRS Project Grant, funded by Cure Cancer Australia and the CanToo Foundation (#1138084), NHMRC Project Grants (APP1158083 and APP1109286), QIMR Berghofer Medical Research Institute Near Miss Funding and a special purpose donation from gratefully received from Sarah Stork.

**Conflict of Interest Statement:** UM has stocks in Abcodia awarded to her by UCL. All other authors declare no potential conflicts of interest.

### Competing Interest Statement

Usha Menon has stocks in Abcodia awarded to her by UCL. All other authors declare no potential conflicts of interest.

### Funding Statement

TAO’M is supported by a National Health and Medical Research Council (NHMRC) Early Career Fellowship (APP1111246) and Investigator Fellowship (APP1173170), ABS is supported by an NHMRC Senior Research Fellowship (APP1061779) and Investigator Fellowship (APP1177524). This work was supported by a Cancer Australia PdCCRS Project Grant, funded by Cure Cancer Australia and the CanToo Foundation (#1138084), NHMRC Project Grants (APP1158083 and APP1109286), QIMR Berghofer Medical Research Institute Near Miss Funding and a special purpose donation from gratefully received from Sarah Stork. Full ECAC and OCAC funding information can be found in the Supplementary Note.

## References

1 Bray, F. et al. Global cancer statistics 2018: GLOBOCAN estimates of incidence and mortality worldwide for 36 cancers in 185 countries. CA Cancer J Clin 68, 394–424, doi:10.3322/caac.21492 (2018).

2 Koshiyama, M., Matsumura, N. & Konishi, I. Recent concepts of ovarian carcinogenesis: type I and type II. Biomed Res Int 2014, 934261, doi:10.1155/2014/934261 (2014).

3 Cramer, D. W. The epidemiology of endometrial and ovarian cancer. Hematol Oncol Clin North Am 26, 1–12, doi:10.1016/j.hoc.2011.10.009 (2012).

4 Braem, M. G. et al. Reproductive and hormonal factors in association with ovarian cancer in the Netherlands cohort study. Am J Epidemiol 172, 1181–1189, doi:10.1093/aje/kwq264 (2010).

5 Dossus, L. et al. Reproductive risk factors and endometrial cancer: the European Prospective Investigation into Cancer and Nutrition. Int J Cancer 127, 442–451, doi:10.1002/ijc.25050 (2010).

6 Gong, T. T., Wang, Y. L. & Ma, X. X. Age at menarche and endometrial cancer risk: a dose-response meta-analysis of prospective studies. Sci Rep 5, 14051, doi:10.1038/srep14051 (2015).

7 Gong, T. T., Wu, Q. J., Vogtmann, E., Lin, B. & Wang, Y. L. Age at menarche and risk of ovarian cancer: a meta-analysis of epidemiological studies. Int J Cancer 132, 2894–2900, doi:10.1002/ijc.27952 (2013).

8 Danforth, K. N. et al. Breastfeeding and risk of ovarian cancer in two prospective cohorts. Cancer Causes Control 18, 517–523, doi:10.1007/s10552-007-0130-2 (2007).

9 Jordan, S. J. et al. Breastfeeding and Endometrial Cancer Risk: An Analysis From the Epidemiology of Endometrial Cancer Consortium. Obstet Gynecol 129, 1059–1067, doi:10.1097/AOG.0000000000002057 (2017).

10 Collaborative Group on Epidemiological Studies of Ovarian, C. a. n. c. e. r. et al. Ovarian cancer and oral contraceptives: collaborative reanalysis of data from 45 epidemiological studies including 23,257 women with ovarian cancer and 87,303 controls. Lancet 371, 303–314, doi:10.1016/S0140-6736(08)60167-1 (2008).

11 Maxwell, G. L. et al. Progestin and estrogen potency of combination oral contraceptives and endometrial cancer risk. Gynecol Oncol 103, 535–540, doi: 10.1016/j.ygyno.2006.03.046 (2006).

12 Jenabi, E. & Poorolajal, J. The effect of body mass index on endometrial cancer: a meta-analysis. Public Health 129, 872–880, doi:10.1016/j.puhe.2015.04.017 (2015).

13 Leitzmann, M. F. et al. Body mass index and risk of ovarian cancer. Cancer 115, 812-822, doi:10.1002/cncr.24086 (2009).

14 Vang, R. et al. Molecular Alterations of TP53 are a Defining Feature of Ovarian High-Grade Serous Carcinoma: A Rereview of Cases Lacking TP53 Mutations in The Cancer Genome Atlas Ovarian Study. Int J Gynecol Pathol 35, 48–55, doi:10.1097/PGP.0000000000000207 (2016).

15 Schultheis, A. M. et al. TP53 Mutational Spectrum in Endometrioid and Serous Endometrial Cancers. Int J Gynecol Pathol 35, 289–300, doi:10.1097/PGP.0000000000000243 (2016).

16 McConechy, M. K. et al. Ovarian and endometrial endometrioid carcinomas have distinct CTNNB1 and PTEN mutation profiles. Mod Pathol 27, 128–134, doi:10.1038/modpathol.2013.107 (2014).

17 Kolbe, D. L. et al. Differential analysis of ovarian and endometrial cancers identifies a methylator phenotype. PLoS One 7, e32941, doi:10.1371/journal.pone.0032941 (2012).

18 Zorn, K. K. et al. Gene expression profiles of serous, endometrioid, and clear cell subtypes of ovarian and endometrial cancer. Clin Cancer Res 11, 6422–6430, doi:10.1158/1078-0432.CCR-05-0508 (2005).

19 King, C. M., Barbara, C., Prentice, A., Brenton, J. D. & Charnock-Jones, D. S. Models of endometriosis and their utility in studying progression to ovarian clear cell carcinoma. J Pathol 238, 185–196, doi:10.1002/path.4657 (2016).

20 Lu, K. H., & Broaddus, R. R. Gynecologic Cancers in Lynch Syndrome/HNPCC. Fam Cancer 4, 249–254, doi:10.1007/s10689-005-1838-3 (2005).

21 O’Mara, T. A. et al. Identification of nine new susceptibility loci for endometrial cancer. Nat Commun 9, 3166, doi:10.1038/s41467-018-05427-7 (2018).

22 Phelan, C. M. et al. Identification of 12 new susceptibility loci for different histotypes of epithelial ovarian cancer. Nat Genet 49, 680–691, doi:10.1038/ng.3826 (2017).

23 Cheng, T. H. et al. Meta-analysis of genome-wide association studies identifies common susceptibility polymorphisms for colorectal and endometrial cancer near SH2B3 and TSHZ1. Sci Rep 5, 17369, doi:10.1038/srep17369 (2015).

24 Kar, S. P. et al. Genome-Wide Meta-Analyses of Breast, Ovarian, and Prostate Cancer Association Studies Identify Multiple New Susceptibility Loci Shared by at Least Two Cancer Types. Cancer Discov 6, 1052–1067, doi:10.1158/2159-8290.CD-15-1227 (2016).

25 Bulik-Sullivan, B. K. et al. LD Score regression distinguishes confounding from polygenicity in genome-wide association studies. Nat Genet 47, 291–295, doi:10.1038/ng.3211 (2015).

26 Turley, P. et al. Multi-trait analysis of genome-wide association summary statistics using MTAG. Nat Genet 50, 229–237, doi:10.1038/s41588-017-0009-4 (2018).

27 Han, B. & Eskin, E. Interpreting meta-analyses of genome-wide association studies. PLoS Genet 8, e1002555, doi:10.1371/journal.pgen.1002555 (2012).

28 Han, B. & Eskin, E. Random-effects model aimed at discovering associations in metaanalysis of genome-wide association studies. Am J Hum Genet 88, 586–598, doi:10.1016/j.ajhg.2011.04.014 (2011).

29 Pickrell, J. K. et al. Detection and interpretation of shared genetic influences on 42 human traits. Nat Genet 48, 709–717, doi:10.1038/ng.3570 (2016).

30 Lawrenson, K. et al. In vitro three-dimensional modelling of human ovarian surface epithelial cells. Cell Prolif 42, 385–393, doi:10.1111/j.1365-2184.2009.00604.x (2009).

31 O’Mara, T. A., Spurdle, A. B., Glubb, D. M. & Endometrial Cancer Association, C. Analysis of Promoter-Associated Chromatin Interactions Reveals Biologically Relevant Candidate Target Genes at Endometrial Cancer Risk Loci. Cancers (Basel) 11, doi:10.3390/cancers11101440 (2019).

32 Servant, N. et al. HiC-Pro: an optimized and flexible pipeline for Hi-C data processing. Genome Biol 16, 259, doi:10.1186/s13059-015-0831-x (2015).

33 Lareau, C. A. & Aryee, M. J. hichipper: a preprocessing pipeline for calling DNA loops from HiChIP data. Nature methods 15, 155–156, doi:10.1038/nmeth.4583 (2018).

34 Phanstiel, D. H., Boyle, A. P., Heidari, N. & Snyder, M. P. Mango: a bias-correcting ChIA-PET analysis pipeline. Bioinformatics 31, 3092–3098, doi:10.1093/bioinformatics/btv336 (2015).

35 Lim, Y. W. et al. Germline genetic polymorphisms influence tumor gene expression and immune cell infiltration. Proc Natl Acad Sci USA 115, E11701-E11710, doi:10.1073/pnas.1804506115 (2018).

36 GTEx, C. o. n. s. o. r. t. i. u. m. The Genotype-Tissue Expression (GTEx) project. Nat Genet 45, 580–585, doi:10.1038/ng.2653 (2013).

37 Võsa, U. et al. Unraveling the polygenic architecture of complex traits using blood eQTL metaanalysis. *bioRxiv*, 447367, doi:10.1101/447367 (2018).

38 Carvajal-Carmona, L. G. et al. Candidate locus analysis of the TERT-CLPTM1L cancer risk region on chromosome 5p15 identifies multiple independent variants associated with endometrial cancer risk. Hum Genet 134, 231–245, doi:10.1007/s00439-014-1515-4 (2015).

39 Chung, C. C., Magalhaes, W. C., Gonzalez-Bosquet, J. & Chanock, S. J. Genome-wide association studies in cancer--current and future directions. Carcinogenesis 31, 111–120, doi:10.1093/carcin/bgp273 (2010).

40 Bojesen, S. E. et al. Multiple independent variants at the TERT locus are associated with telomere length and risks of breast and ovarian cancer. Nat Genet 45, 371–384, 384e371–372, doi:10.1038/ng.2566 (2013).

41 Yuan, X., Larsson, C. & Xu, D. Mechanisms underlying the activation of TERT transcription and telomerase activity in human cancer: old actors and new players. Oncogene 38, 6172–6183, doi:10.1038/s41388-019-0872-9 (2019).

42 Pestana, A., Vinagre, J., Sobrinho-Simoes, M. & Soares, P. TERT biology and function in cancer: beyond immortalisation. J Mol Endocrinol 58, R129-R146, doi:10.1530/JME-16-0195 (2017).

43 Bailey, M. H. et al. Comprehensive Characterization of Cancer Driver Genes and Mutations. Cell 173, 371–385 e318, doi:10.1016/j.cell.2018.02.060 (2018).

44 Tashiro, E., Tsuchiya, A. & Imoto, M. Functions of cyclin D1 as an oncogene and regulation of cyclin D1 expression. Cancer Sci 98, 629–635, doi:10.1111/j.1349-7006.2007.00449.x (2007).

45 Berger, A. C. et al. A Comprehensive Pan-Cancer Molecular Study of Gynecologic and Breast Cancers. Cancer Cell 33, 690–705 e699, doi:10.1016/j.ccell.2018.03.014 (2018).

46 Painter, J. N. et al. Fine-mapping of the HNF1B multicancer locus identifies candidate variants that mediate endometrial cancer risk. Hum Mol Genet 24, 1478–1492, doi:10.1093/hmg/ddu552 (2015).

47 Spurdle, A. B. et al. Genome-wide association study identifies a common variant associated with risk of endometrial cancer. Nat Genet 43, 451–454, doi:10.1038/ng.812 (2011).

48 Pharoah, P. D. et al. GWAS meta-analysis and replication identifies three new susceptibility loci for ovarian cancer. Nat Genet 45, 362–370, 370e361–362, doi:10.1038/ng.2564 (2013).

49 Shen, H. et al. Epigenetic analysis leads to identification of HNF1B as a subtype-specific susceptibility gene for ovarian cancer. Nat Commun 4, 1628, doi:10.1038/ncomms2629 (2013).

50 O’Mara, T. A., Glubb, D. M., Kho, P. F., Thompson, D. J. & Spurdle, A. B. Genome-Wide Association Studies of Endometrial Cancer: Latest Developments and Future Directions. Cancer Epidemiol Biomarkers Prev 28, 1095–1102, doi:10.1158/1055-9965.EPI-18-1031 (2019).

51 Pearce, C. L. et al. Association between endometriosis and risk of histological subtypes of ovarian cancer: a pooled analysis of case-control studies. Lancet Oncol 13, 385–394, doi:10.1016/S1470-2045(11)70404-1 (2012).

52 Painter, J. N. et al. Genetic overlap between endometriosis and endometrial cancer: evidence from cross-disease genetic correlation and GWAS meta-analyses. Cancer Med 7, 1978–1987, doi:10.1002/cam4.1445 (2018).

53 Nilufer, R. et al. Large-scale genome-wide association meta-analysis of endometriosis reveals 13 novel loci and genetically-associated comorbidity with other pain conditions. *bioRxiv* (2018).

54 Chen, C. L. et al. Stat3 activation in human endometrial and cervical cancers. Br J Cancer 96, 591–599, doi:10.1038/sj.bjc.6603597 (2007).

55 Yoshikawa, T. et al. JAK2/STAT3 pathway as a therapeutic target in ovarian cancers. Oncol Lett 15, 5772–5780, doi:10.3892/ol.2018.8028 (2018).

56 Khaled, W. T. et al. BCL11A is a triple-negative breast cancer gene with critical functions in stem and progenitor cells. Nat Commun 6, 5987, doi:10.1038/ncomms6987 (2015).

57 Lazarus, K. A. et al. BCL11A interacts with SOX2 to control the expression of epigenetic regulators in lung squamous carcinoma. Nat Commun 9, 3327, doi:10.1038/s41467-018-05790-5 (2018).

58 Luc, S. et al. Bcl11a Deficiency Leads to Hematopoietic Stem Cell Defects with an Aging-like Phenotype. Cell Rep 16, 3181–3194, doi:10.1016/j.celrep.2016.08.064 (2016).

59 Itamochi, H. et al. Whole-genome sequencing revealed novel prognostic biomarkers and promising targets for therapy of ovarian clear cell carcinoma. Br J Cancer 117, 717–724, doi:10.1038/bjc.2017.228 (2017).

60 Er, T. K. et al. Targeted next-generation sequencing for molecular diagnosis of endometriosis-associated ovarian cancer. J Mol Med (Berl) 94, 835–847, doi:10.1007/s00109-016-1395-2 (2016).

61 Arend, P. Position of human blood group O(H) and phenotype-determining enzymes in growth and infectious disease. Ann N Y Acad Sci, doi:10.1111/nyas.13694 (2018).

62 Meyer, C. et al. The MLL recombinome of acute leukemias in 2013. Leukemia 27, 2165–2176, doi:10.1038/leu.2013.135 (2013).

63 Nakazawa, K., Dashzeveg, N. & Yoshida, K. Tumor suppressor p53 induces miR- 1915 processing to inhibit Bcl-2 in the apoptotic response to DNA damage. FEBS J 281, 2937–2944, doi:10.1111/febs.12831 (2014).

64 Ruth, K. S. et al. Genome-wide association study with 1000 genomes imputation identifies signals for nine sex hormone-related phenotypes. Eur J Hum Genet 24, 284-290, doi:10.1038/ejhg.2015.102 (2016).

65 Domanski, T. L., Finta, C., Halpert, J. R. & Zaphiropoulos, P. G. cDNA cloning and initial characterization of CYP3A43, a novel human cytochrome P450. Mol Pharmacol 59, 386–392, doi:10.1124/mol.59.2.386 (2001).

66 Downie, D. et al. Profiling cytochrome P450 expression in ovarian cancer: identification of prognostic markers. Clin Cancer Res 11, 7369–7375, doi:10.1158/1078-0432.CCR-05-0466 (2005).

67 Prossnitz, E. R. & Barton, M. The G-protein-coupled estrogen receptor GPER in health and disease. Nat Rev Endocrinol 7, 715–726, doi:10.1038/nrendo.2011.122 (2011).

68 Clark, B. J., Prough, R. A. & Klinge, C. M. Mechanisms of Action of Dehydroepiandrosterone. Vitam Horm 108, 29–73, doi:10.1016/bs.vh.2018.02.003 (2018).

69 Kuchenbaecker, K. B. et al. Identification of six new susceptibility loci for invasive epithelial ovarian cancer. Nat Genet 47, 164–171, doi:10.1038/ng.3185 (2015).

